# Pathogen- and type-specific changes in invasive bacterial disease epidemiology during the first year of the COVID-19 pandemic in the Netherlands

**DOI:** 10.1101/2022.04.19.22273034

**Authors:** Anneke Steens, Mirjam J. Knol, Wieke Freudenburg-de Graaf, Hester E. de Melker, Arie van der Ende, Nina M. van Sorge

**Author notes:** **Corresponding author:** Prof. dr. N.M. van Sorge, Amsterdam UMC location University of Amsterdam, Department of Medical Microbiology and Infection Prevention, Netherlands Reference Laboratory for Bacterial Meningitis, Meibergdreef 9, 1105 AZ Amsterdam, The Netherlands.

## Abstract

The COVID-19 control measures have resulted in a decline in invasive bacterial disease caused by *Neisseria meningitidis* (IMD), *Streptococcus pneumoniae* (IPD), and *Haemophilus influenzae* (Hi-D). The species comprise different serogroups and serotypes that impact transmissibility and virulence. We evaluated type- and pathogen-specific changes in invasive bacterial disease epidemiology in the Netherlands during the first year of the SARS-CoV-2 pandemic. Cases were based on nationwide surveillance for five bacterial species with either respiratory (IMD, IPD, Hi-D) or non-respiratory (controls) transmission routes and compared between the pre-COVID period (April 2015-March 2020) and the first COVID-19 year (April 2020-March 2021). IMD, IPD, and Hi-D cases decreased by 78%, 67%, and 35%, respectively, in the first COVID-19 year compared to the pre-COVID period although effects differed per age group. Serogroup B-IMD declined by 61%, while serogroup W and Y-IMD decreased >90%. IPD caused by serotypes 7F, 15A, 12F, 33F, and 8 showed the most pronounced decline (≥76%). In contrast to an overall decrease in Hi-D cases, vaccine-preventable serotype b (Hib) increased by 51%. COVID-19 control measures had pathogen- and type-specific effects related to invasive infections. Continued surveillance is critical to monitor potential rebound effects once restriction measures are lifted and transmission is resumed.

## Introduction

Invasive bacterial infections, such as meningitis and sepsis, are associated with high rates of mortality and morbidity worldwide. In the Netherlands as well as globally, the most common causes of bacterial invasive disease differ per age group and availability and implementation of preventive measures ^1^. For meningitis for example, *Streptococcus agalactiae* and *Escherichia coli* cause nearly 64% of all bacterial meningitis cases in infants (0-89 days of age) in the Netherlands^1^, with no vaccines available to prevent these infections. *Neisseria meningitidis, Streptococcus pneumoniae*, and *Haemophilus influenzae* are main causes of invasive disease in children under the age of 5 years old and adolescents (for *N. meningitidis*) ^1-4^.

Nationwide surveillance of invasive bacterial infections caused by aforementioned pathogens allows close monitoring of their incidences and evaluation of the effect of vaccination. In 1993, vaccination with a glycoconjugate vaccine against *H. influenzae* type b (Hib) was implemented in the National Immunisation Programme (NIP)^3^. In 2002, vaccination with a glycoconjugate vaccine against *N. meningitidis* serogroup C (MenC) was introduced at 14 months, and was accompanied by a catch-up campaign for 1-18-year olds. MenC was replaced by MenACYW in 2018 ^5^. In addition, a catch-up campaign among 14-18-year olds with MenACWY was organized and the vaccine is routinely provided to 14-year olds through the NIP since 2020 ^6^. For *S. pneumoniae*, a 7-valent pneumococcal conjugate vaccine (PCV7) was implemented in 2006 and replaced by the 10-valent PCV10 in 2011 ^7^. The implementation of vaccines for specific serotypes of *S. pneumoniae* and *H. influenzae*, and for serogroups of *N. meningitidis* in the routine NIP has dramatically changed the epidemiology of invasive bacterial disease in the last 20-30 years ^1-4^, decreasing the incidence of invasive bacterial disease caused by vaccine-covered serotype- and serogroup-specific pathogens in children by > 90% ^1,7,8^. Additionally, declines of these targeted serogroups and -types have been observed in non-targeted age groups as a result of indirect protection ^2,9^.

The occurrence of the COVID-19 pandemic in 2020 resulted in the implementation of public health response measures such as working from home, social 1.5-meter distancing, and school and day care closures, which varied in stringency and enforcement, from March 15, 2020 onwards ^10^. These control measures did not just limit transmission and subsequent disease caused by SARS-CoV-2, but concomitantly interrupted person-to-person transmission of other pathogens. Indeed, a decline in the incidence of invasive bacterial infections with respiratory transmission including those caused by *N. meningitidis* (IMD), *S. pneumoniae* (IPD) and *H. influenzae* (Hi-D) has been observed in many countries and regions ^11-13^. In contrast, the incidence of neonatal invasive diseases caused by *S. agalactiae* appears to have been unaffected by the COVID-19-related measures ^11^. Indeed, transmission of this pathogen is likely unaffected by control measures as transmission results from vertical instead of respiratory transmission. This is also the case for neonatal invasive disease caused by *E. coli*. Therefore, invasive disease cases due to these (neonatal) pathogens can be used as a control to exclude impact of the pandemic on isolate submission by microbiology laboratories.

Here, we retrospectively analysed changes in the epidemiology of invasive bacterial diseases in the Netherlands during the first year of the SARS-CoV-2 pandemic, with specific focus on differences between bacterial species and subtypes. We compared the number of cases and incidence of invasive disease caused by *N. meningitidis, S. pneumoniae, H. influenzae, S. agalactiae*, and *E. coli* based on nationwide microbiological surveillance from the period April 2015 – March 2021.

## Materials & Methods

### Data collection and patient information

Nationwide surveillance data were obtained from the Netherlands Reference Laboratory for Bacterial Meningitis (NRLBM) for all studied pathogens. The NRLBM receives bacterial isolates from patients with invasive bacterial disease (isolated from blood and/or cerebrospinal fluid, CSF), including IPD for children < 5 years of age, and Hi-D and IMD isolates from patients of all ages. In addition, IPD isolates from patients of all ages are received from nine sentinel laboratories, covering about 25% of the Dutch population. *E. coli* and *S. agalactiae* isolates recovered from blood or cerebrospinal fluid (CSF) are received from patients < 1 years of age. All isolates are typed according to the methods described below.

### Typing of bacterial isolates

*N. meningitidis* serogroup was determined by Ouchterlony gel diffusion ^14^. In case of a culture-negative PCR-positive sample, *N. meningitidis* serogroup was determined by real-time PCR using group-specific probes ^15^. *S. pneumoniae* and *H. influenzae* isolates were serotyped by co-agglutination with specific antisera. For *S. pneumoniae*, additional subtyping was assessed by capsular swelling (Quellung reaction) using specific antisera (Serum Staten Institute, Denmark) ^16^. *E. coli* isolates were tested for expression of K1 antigen by phage typing according to the manufacturer (SSI Diagnostica) and H- and O-types based on whole-genome sequences ^17^. *S. agalactiae* isolates were grouped (Lancefield B) and serotyped through agglutination using specific antisera ^18^.

### Study period and patient categories

For this study, data were extracted from the NRLBM database for the period April 2015 until and including March 2021. The period April 2020 – March 2021 represents the COVID-19 period (first lockdown in the Netherlands started on 15 March 2020). Data from the preceding five years (April 2015 – March 2020) were used as comparison and representative for non-COVID-19 years.

For neonatal/infant invasive disease caused by *E. coli* and *S. agalactiae*, data of infants younger than one year were used, since this is resulting from vertical transmission and/or non-respiratory transmission. For *N. meningitidis, S. pneumoniae*, and *H. influenzae*, patients were categorized into age-groups as appropriate for the epidemiology of the respective pathogens. Specifically, for IPD and Hi-D, age categories included the under-fives, 5-64 years, and older adults (65+). For IMD, the 5-19-year olds and 20-64-year olds were analysed separately in addition to the under-fives and 65+ age groups. For IPD, the age group 73-79 years was excluded from the analyses to avoid a possible bias from the introduction of the PPV23 vaccine in autumn 2020 for this cohort.

IPD data were categorized on vaccine-type, *i*.*e*. PCV10 serotypes (1, 4, 5, 6B, 7F, 9V, 14, 18C, 19F, 23F), PCV13 extra (3, 6A, 19A), PPV23 extra (2, 8, 9N, 10A, 11A, 12F, 15B, 17F, 20, 22F, 33F), and non-vaccine serotypes (all remaining serotypes). Non-typeable isolates and isolates with missing serotype information were excluded from the serotype-specific analyses. Specifically, this applied for 0.7% missing or non-typeable serotypes for *S. pneumoniae*, 0.9% missing or ungroupable serogroups for *N. meningitidis*, 0.1% missing serotypes for *H. influenzae*.

### Descriptive analyses

Analyses were performed separately for the different pathogens and serotypes/serogroups. The percentage change was determined as 1-(the number of cases in the COVID-19 year / the average number of cases in the five preceding years)*100. The 5-year moving average per month for the pre-COVID period was based on the targeted month, one month before and one after the targeted month. The incidence was determined by dividing the number of cases in the defined years (April-March) by the number of inhabitants for the same period (for a specific age group) / 100,000. Population statistics were obtained from Statistics Netherlands. The percentage change per serotype/serogroup was determined relative to the number of isolates with known serotype/serogroup. For IPD, results on serotype-specific changes are presented if the serotype had a yearly average of at least 10 cases in the five pre-COVID-19 years. Ranking was used to determine whether serotypes increased/decreased differentially over time.

### Ethical statement

In accordance with Dutch law, approval from a medical ethics committee was not deemed necessary since cases were not subject to any actions or rules of conduct. Data regarding cases were obtained by use of standard surveillance procedures, and pseudonymised data were used in the study. Informed consent was not obtained, as the collection of data complies with the exceptions for not asking informed consent as formulated in the Dutch Implementation Act General Data Protection Regulation.

## Results

### Decrease of Invasive disease cases caused by respiratory-transmitted pathogens in the first COVID-19 year

Overall, IMD, IPD, and Hi-D decreased by 78% (n=36 versus 163), 67% (n=175 versus 531) and 35% (n=144 versus 221) compared to the average number of cases in the preceding five years 2015-2020 (pre-COVID), respectively. The decrease was immediately apparent in April 2020, which is the first month following lock-down (15 March 2020), for all three pathogens (Figure 1A-C). IMD and IPD case numbers remained lower compared to non-COVID years all throughout the first COVID-19 year (Figure 1A, B). In contrast, Hi-D case numbers were similar or higher compared to the 5-year moving average in August, October, and December 2020 (Figure 1C). Invasive disease cases caused by *S. agalactiae* and *E. coli* showed an increase in the 2015-2020 period, with similar or increased number of cases in the first COVID-19 year (n=131 versus 82 on average and n=80 versus 50 on average, respectively; Figure 1D, E).

**Figure 1.**
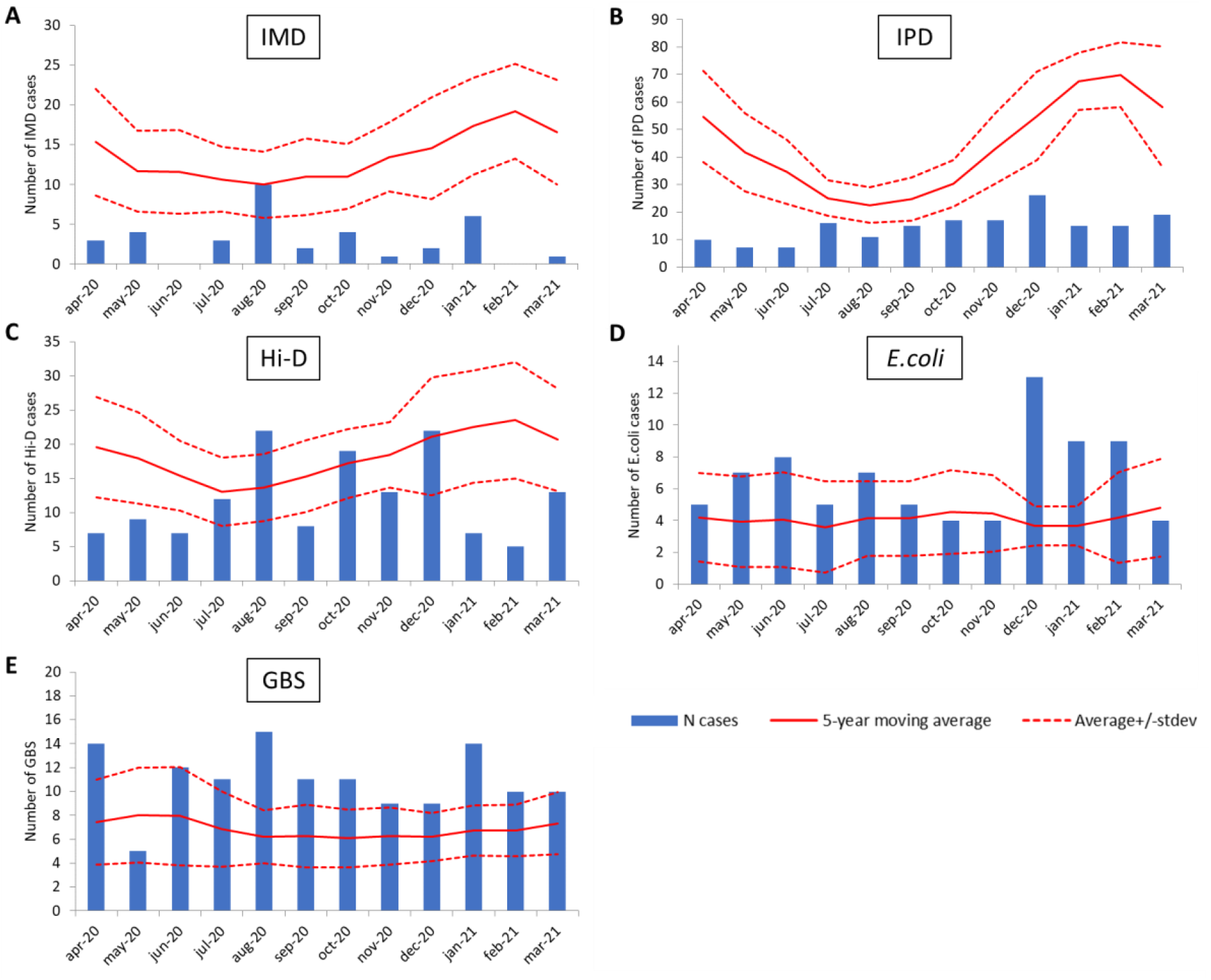
Number of invasive disease cases by month in the first COVID-19 year (2020-2021; bars) based on bacteriological surveillance, the 5-year moving average in pre-COVID years (solid line) and its standard deviation for the pre-COVID period (dashed line) for A: N. meningitidis (IMD), B: S. pneumoniae (IPD), C: H. influenzae (Hi-D), D: E. coli, E: S. agalactiae. Note that the y-axis differs between pathogens.

### Age-specific effects on epidemiology of invasive disease

We assessed whether the reduction in invasive disease cases was similar across different age groups despite small numbers. For IMD and IPD, the reduction in invasive disease cases was observed across the different age categories studied; IMD and IPD cases were 58% (n=8 versus 17) and 53% (n=13 versus 31 pre-COVID) lower for children under the age of five years, and 86% (n=6 versus 43) and 69% (n=89 versus 283) lower in the 65+ population compared to the pre-COVID average. In contrast, Hi-D cases increased by 13% (n=38 versus 34) in the under-fives but decreased by 18% (n=63 versus 77) in 5-64-year olds and 61% in the 65+ age group (n=42 versus 109). These age-specific differences can be explained by serotype-specific effects, as presented below.

### Serogroup-specific changes in invasive meningococcal disease cases

Of the 851 *N. meningitidis* isolates received during the study period, 397 (47%) were serogroup B, 29 (3.4%) serogroup C, 313 (37%) serogroup W and 100 (11.8%) serogroup Y. Serogroup A-IMD did not occur in the study period. Serogroup W-IMD and Y-IMD cases declined by 92% (5 versus 62 on average) and 90% (2 versus 20 on average), respectively, in the first COVID-19-year compared to the average number of cases in the previous 5 years (Figure 2B, C). These case numbers corresponded to incidences of 0.03/100,000 for serogroup W-IMD and 0.02/100,000 for serogroup Y-IMD in 2020-2021. The decline in serogroup B-IMD cases was less pronounced than for serogroups W and Y, with 62% reduction in cases (n=28 versus 74 on average; Figure 2A), resulting in an incidence of 0.16/100,000 compared to 0.38-0.48/100,000 in the pre-COVID-19 period. Serogroup C-IMD was not detected during the first COVID-19-year.

**Figure 2.**
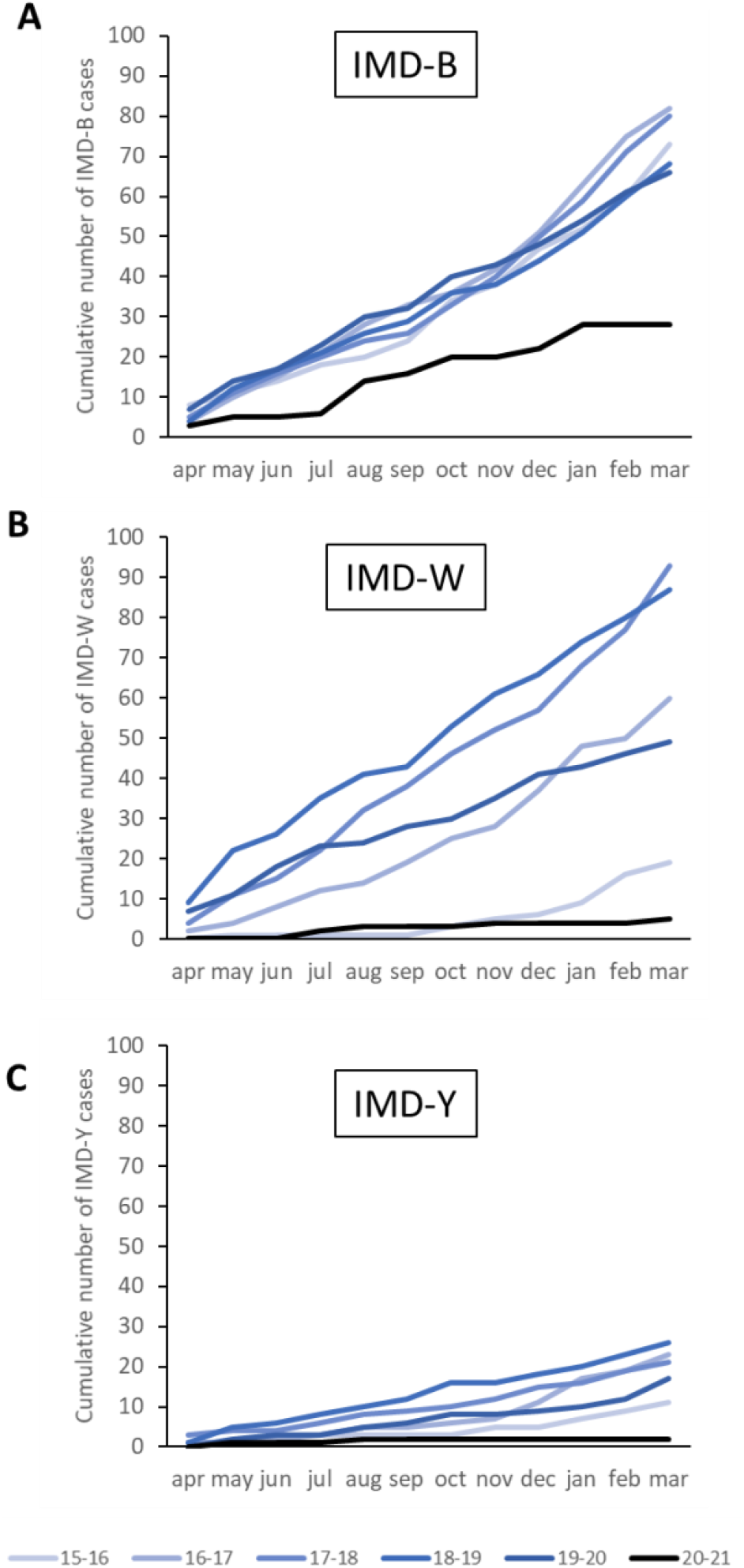
Cumulative number of cases of invasive meningococcal disease caused by A: serogroup B, B: serogroup W and C: serogroup Y in the first COVID-19 year (2020-2021; black) compared to the previous five pre-COVID years (blue).

### Serotype-specific changes in invasive pneumococcal disease cases

Overall, 2,830 IPD cases were included in the study. In the five pre-COVID-19 years, an average of 531 IPD isolates per year were submitted by the sentinel laboratories, resulting in an overall incidence of 12.3-14.3/100,000. In the first COVID-19 year, the number of IPD cases decreased to 175 cases (67% reduction) resulting in an overall incidence of 4.3/100,000 among all age groups excluding the 73-79-year olds. The proportion of PCV10 serotypes declined over the study period (Figure 3), possibly related to the implementation of PCV10 in the NIP since 2011. Dissection of the non-PCV10 serotypes in the first COVID-19 year showed a reduction of IPD cases caused by additional serotypes covered by PCV13 (PCV13 extra) of 55% (n=56 versus 124 on average) as well as non-PPV23 serotypes of 53% (n=43 versus 92 on average) (Figure 3). In comparison, IPD cases caused by additional PPV23 (PPV23extra) serotypes decreased by 74% (n=69 versus 261 on average; Figure 3). As a result, the proportion of PCV13extra serotypes and that of non-PPV23 serotypes increased (Figure 3).

**Figure 3:**
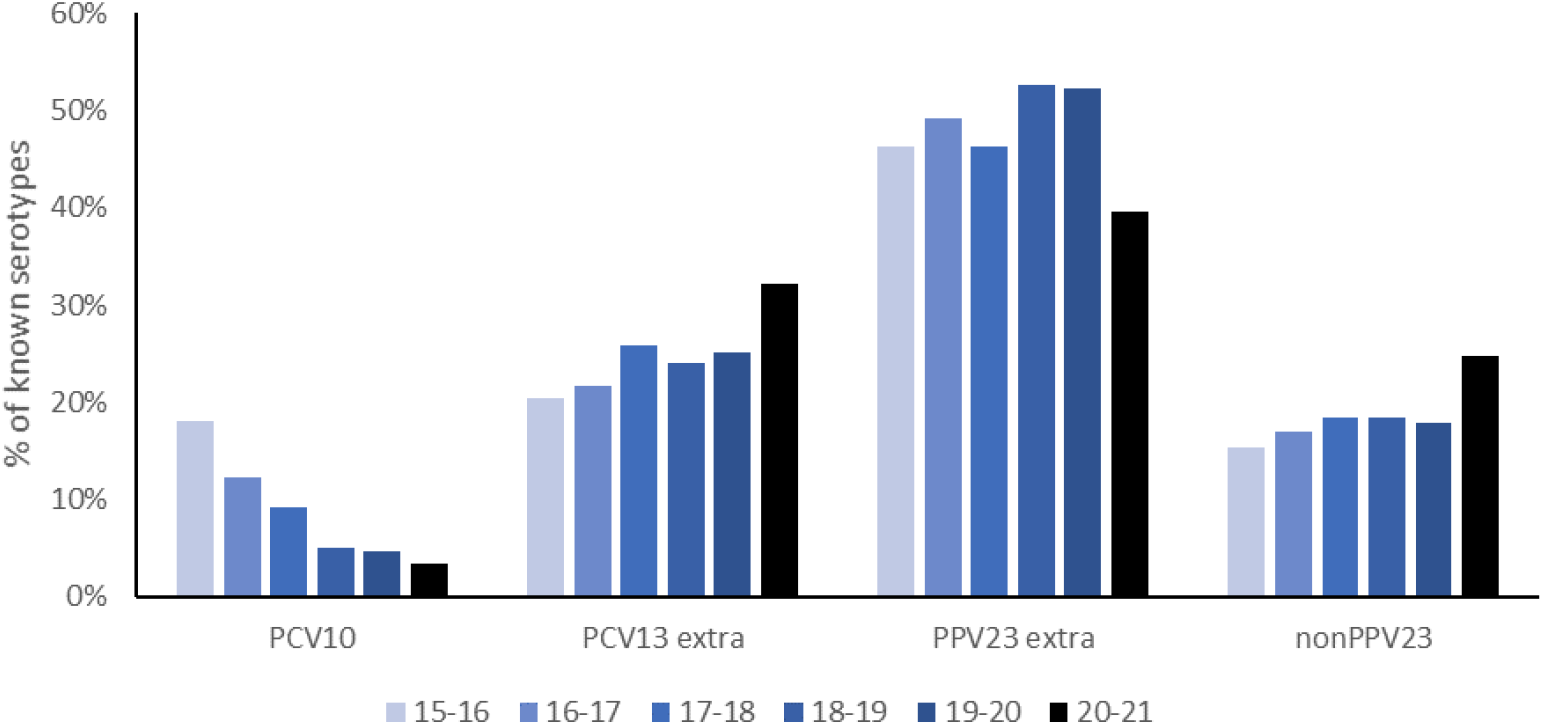
Percentage of IPD cases caused by vaccine serotypes during the years 2015-2021, in all age groups except those aged 73-79 years old.

We also analysed changes in the number of IPD cases caused by individual pneumococcal serotypes (Table 1). Only serotypes that changed by more than 10% compared to the average change of all-serotype IPD (67%) are discussed. The proportional increase among PCV13extra serotypes was mainly attributable to serotype 19A, which was less affected (50% decrease) than the average IPD decline (67%; Table 1). For PPV23 serotypes, a larger than average decrease was observed for serotypes 8 (76% decrease), 12F (82%), and 33F (87%), as well as non-vaccine serotype 15A (86%; Table 1). Of the non-vaccine serotypes, serotype 6C showed the smallest reduction, decreasing by only 16% compared to the average proportion in the previous five years (Table 1). Note, however, that the ranking of the most common serotypes remained stable, with serotypes 19A and 8 being the two most common serotypes for all analysed years and serotypes 3, 7F, 22F and 9N remaining in the top 3-6 rank.

**Table 1.** Number of cases of invasive pneumococcal disease per serotype as average and range of pre-COVID period (2015-2020) and first COVID-19 year in all age groups except those aged 73-79 years old. Decrease in number of serotype-specific IPD cases is indicated by percentage. * If covered by PCV10, then PCV10 is used, if not in PCV10 but in PCV13, then PCV13 is used, if not in PCV13 and PCV10 but in PPV23, then PCV23 is used. If not covered by any of the currently available vaccines, non-vaccine type (NVT) is used.

| Serotype | Covered by vaccine * | Average annual number (range) of cases, April 2015-March 2020 | Number of cases in first COVID year (range), April 2021 – March 2021 | Decrease in COVID versus non-COVID year (%) |
| --- | --- | --- | --- | --- |
| All serotypes |  | 531 (503-576) | 175 | 67% |
| 8 | PPV23 | 126 (118-129) | 30 | 76% |
| 19A | PCV13 | 76 (73-83) | 38 | 50% |
| 3 | PCV13 | 46 (38-59) | 18 | 61% |
| 22F | PPV23 | 35 (27-39) | 11 | 68% |
| 9N | PPV23 | 28 (21-34) | 9 | 68% |
| 12F | PPV23 | 23 (19-27) | 4 | 82% |
| 7F | PCV10 | 21 (4-43) | 0 | 100% |
| 6C | NVT | 18 (16-22) | 15 | 16% |
| 33F | PPV23 | 16 (11-20) | 2 | 87% |
| 15A | NVT | 14 (12-15) | 2 | 86% |
| 23B | NVT | 13 (10-16) | 5 | 60% |
| 23A | NVT | 11 (6-15) | 4 | 64% |
| 1 | PCV10 | 11 (0-30) | 0 | 100% |
| 10A | PPV23 | 11 (8-14) | 4 | 62% |

#### Serotype-specific changes in *H. influenzae* invasive disease

*H. influenzae* can be categorized in encapsulated (type a, b, c, d, e, f) and non-encapsulated strains, also called non-typeable *H. influenzae* (NTHi). The introduction of glycoconjugate vaccines against Hib in 1993 has resulted in a decline of Hib cases, while the number of invasive disease cases caused by NTHi increased in pre-COVID times ^19^. We have focused our analysis on Hib and NTHi, since they cover 90% of all Hi-D cases. Overall, compared to the five preceding years, the number of Hi-D cases decreased by 35% (144 versus 221 on average) in the first COVID year (Figure 1C). However, there was great disparity between Hib and NTHi cases. The number of Hib cases increased by 51% (65 versus 43 on average) (Figure 4A)^19^, resulting in an incidence of 0.37/100,000 in the first COVID-19 year compared to 0.21-0.29/100,000 in the previous 5-year period. This increase was most pronounced in the age group 5-64 years (97% increase; n=26), followed by children < 5 years of age (42%; n=25) and 65+ year-olds (29%; n=15). In contrast, compared to the pre-COVID period, NTHi cases declined by 57% (n=67 versus 154 on average) (Figure 4B), lowering the incidence from 0.73-1.01/100,000 to 0.38/100,000. During the first COVID-19 year, the number of NTHi cases was significantly lower than the 5-year moving average during all months except in August (Figure 4 B), whereas Hib cases were higher than the 5-year moving average for nearly all months (Figure 4 A).

**Figure 4.**
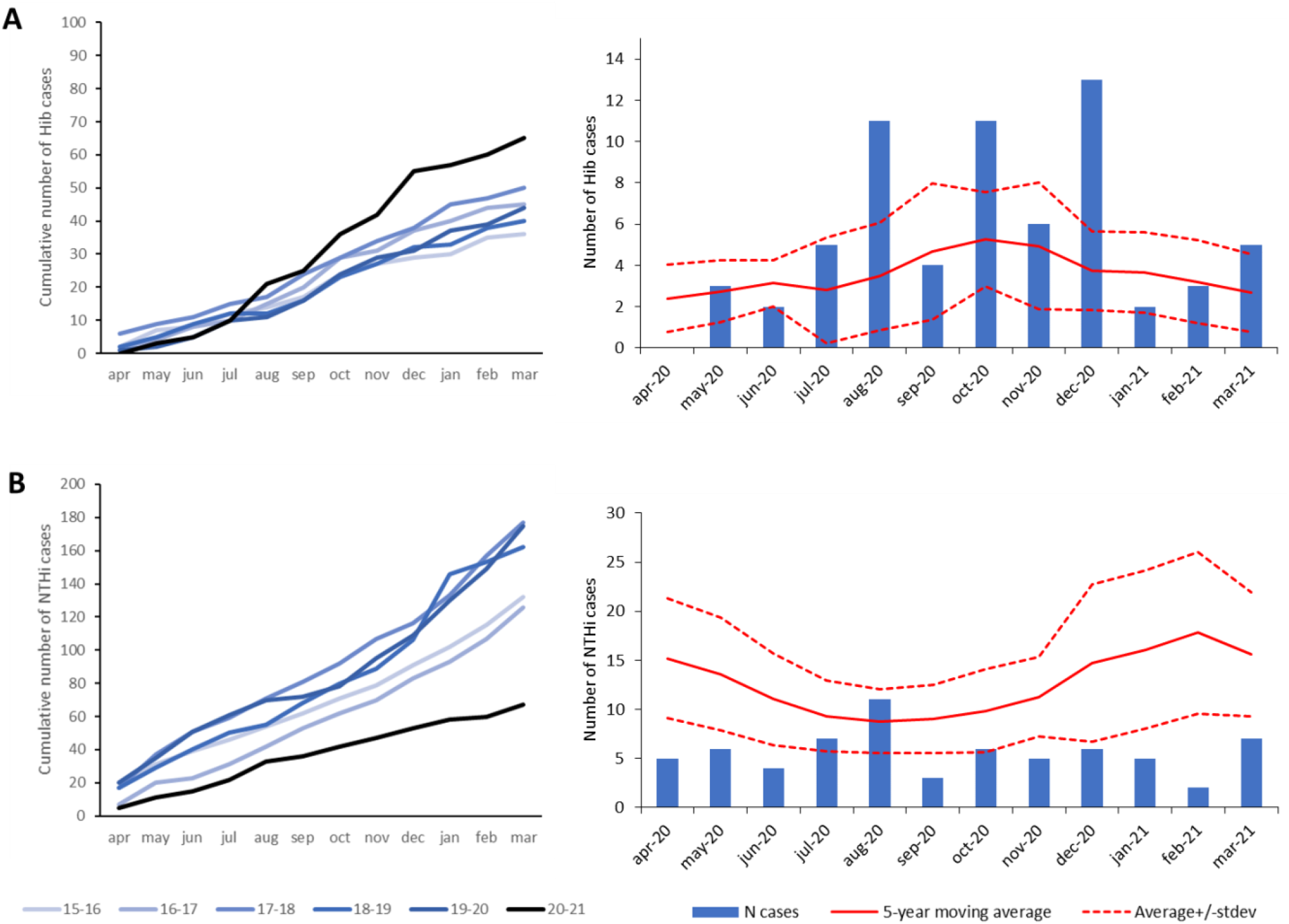
Cumulative number of cases (left) and monthly number of cases (right) for the first COVID-19 year (bars) compared to the 5-year moving average in pre-COVID years (solid line) and the moving average plus/minus its standard deviation for the pre-COVID period (dashed line) for, A: Haemophilus influenzae serotype b (Hib) and B: non-typeable H. influenzae (NTHi).

## Discussion

In the present study, we evaluated the effect of the implementation of public health measures in response to the COVID-19 pandemic by analysing epidemiological changes in the occurrence of invasive disease caused by different bacterial pathogens. We observed a 35% to 78% reduction in the incidence of invasive disease caused by *N. meningitidis, S. pneumoniae* and *H. influenzae*. In contrast, no decrease but a slight increase in infant invasive disease cases caused by *S. agalactiae* and *E. coli* was observed in the COVID-19-year compared to the 2015-2020 pre-COVID period, which is consistent with the observed trend already over a longer period. This suggests that national surveillance, which relies on voluntary isolate submission from clinical diagnostics laboratories, was not generally disrupted. However, we cannot fully exclude that surveillance was affected by care and mass diagnostics of COVID-19 patients or that specific age groups were differentially affected as we were not able to estimate possible age-specific changes in our control group. Because of the surveillance system itself, with subtyping being done by the reference laboratory after isolate submission, it is unlikely that potential effects on surveillance would differ between sub-types.^20^.

A decrease in occurrence of IMD during the COVID-19 pandemic has been observed by many countries ^11,13^. For the results presented here, it is important to note that the MenACWY vaccine has been implemented in the Netherlands in 2018 ^5^. Consequently, the decrease in IMD cases caused by serotypes W and Y are partially influenced by the protective effect from MenACWY vaccination ^6^. The decrease in MenB cases by 62% therefore most accurately reflects the impact of COVID-19 on IMD, since IMD-B disease is not affected by a recent introduction of a vaccine.

Changes in IPD were different for different serotypes. Specifically, serotypes 8, 12F, 33F (all in PPV23 but not in PCV10) and 15A (non-vaccine type) decreased more than other serotypes. In Taiwan, a similarly large decrease in serotype 15A was observed ^21^. Serotypes 8, 12F, and 15A have been increasing in Europe before the COVID-19 pandemic and are known to have a high invasive capacity ^22-26^. Generally, these serotypes are dominant in younger individuals and those without medical underlying conditions ^22^. Unfortunately, we lack information on underlying medical risk conditions in the study cohort. In contrast to the abovementioned serotypes, serotypes 19A and 6C showed a smaller decrease among IPD cases. This is of interest since these serotypes have become more common in pre-COVID years in several countries that use PCV10 ^27^, likely as a result of serotype replacement ^28^. Whether the relatively limited decrease can be partly explained by an offset caused by serotype replacement after PCV10 introduction is yet unknown ^29^.

Our serotype-specific observations differ from reports by some other countries. In Germany, the proportion of vaccine serotypes (PCV13, PCV15, PCV20) remained constant during the COVID-19 control measures, although 12F and 22F seemed to represent a smaller proportion in 2021 than in earlier years indicating a larger decrease during COVID-times and “other serotypes” comprised a larger proportion in 2021 ^30^. In Taiwan, they noticed a large decrease in 19A in contrast to our modest reduction in this serotype ^21^. Finally, in Switzerland, no serotype-specific patterns were observed during the period with COVID-19 control measures, although serotype 23B increased more than other serotypes after easing the non-pharmaceutical interventions ^29^. Serotype-specific changes may reflect age-specific (adherence to) non-pharmaceutical interventions. Furthermore, the serotype-specific changes may reflect differences in propensity of these serotypes to cause secondary infections following respiratory viral infections. Indeed, there were large decreases in overall viral infections during the COVID-19 period, while carriage patterns were reported not to be affected during the COVID-19 pandemic ^31^.

For Hi-D, there was a discrepancy in the changes during the COVID-19 year, which were attributed to different behaviour of serotypes. NTHi disease cases decreased across all age groups, whereas Hib increased in under-fives and 5-64-year olds (specifically in 50-64-year olds), but decreased in the 65+ population. The Hib increase during the first COVID-19 year is striking as preventive measures against COVID-19 were in place and diseases caused by other respiratory-transmitted pathogens decreased. Although not the focus of this study, the vaccine effectiveness against Hib disease did not seem to be different from previous years ^19^. We are currently investigating the possible cause of this increase in Hib disease.

Overall, we showed that the changes in epidemiology during the first COVID-19 year differed between pathogens, subtypes and age groups, with the largest discrepancy seen for Hib and NTHi. Our study indicates that overall the disease burden of respiratory bacterial pathogens was decreased by the preventive measures against COVID-19, but it also shows that other factors affected the epidemiology simultaneously; for IMD, the recent MenACWY-vaccination campaign likely played a role in the observed differences between serogroup B versus serogroup W and Y IMD. For Hib, unknown factors seem to have affected the epidemiology ^19^. The differences in pneumococcal serotype-specific changes might be associated with the epidemic potential, invasive capacity, and/or propensity of the different serotypes to cause secondary infections. Possible differences by risk-group and age in (adherence to) control measures may have played a role in causing different decreases for different age groups. Altogether, we stress the importance of subtype-specific surveillance, also during the COVID-19 pandemic and specifically when measures are relaxed again, given the worry that the interruption of transmission has created a larger group of susceptible individuals ^32^.

## Supporting information

IRB decision on study ethics

## Data Availability

The data presented in the study are available on request with the corresponding author but are not publicly available due to regulations in the Personal Data Protection Act.

## Author contributions

NvS and AvdE were involved in data acquisition. All authors were involved in data analysis and interpretation. NvS prepared the original draft of the manuscript, which was reviewed and edited by all authors. All authors have approved the submitted version of the manuscript.

