## Supplementary material for "Pathogen- and type-specific changes in invasive bacterial disease epidemiology during the first year of the COVID-19 pandemic in the Netherlands": IRB decision on study ethics

Rijksinstituut voor Volksgezondheid
en Milieu
*Ministerie van Volksgezondheid,*
*Welzijn en Sport*

IIV/KIM

A. van Leeuwenhoeklaan 9
3721 MA Bilthoven
Postbus 1
3720 BA Bilthoven
www.rivm.nl

mensgebonden-


**Auteur**
Dr. R. Bos

T 088 689 3439

[redacted]

| | | |
|---|---|---|
| Datum | : | 13 april 2022 |
| Onderzoeker(s) | : | Anneke Steens |
| Titel onderzoek | : | Type/specific changes in invasive bacterial diseases during COVID |
| RIVM studienummer | : | EPI-560 |

[redacted] mevr. Steens,

*English summary: At your request, the Centre for Clinical Expertise at the RIVM assessed the above-mentioned research proposal. We verified whether the work complies with the specific conditions as stated in the law for medical research involving human subjects (WMO). We are of the opinion that the research does <u>not</u> fulfill one or both of these conditions and therefore conclude it is exempted for further approval by the ethical research committee.*

[redacted]

[redacted]

[redacted]

[redacted],

Dhr Dr. R. (Rogier) Bos, hoofd KIM

[redacted]
